# Mental Stress-Induced Myocardial Ischemia and Cardiovascular Events in Women after a Premature Myocardial Infarction: A Prospective Cohort Study

**DOI:** 10.64898/2026.09.30.26364456

**Authors:** Viola Vaccarino, Amit J. Shah, Hua She, Lisa Elon, Tené T. Lewis, Yi-An Ko, Nancy Murrah, Lucy H. Shallenberger, Tatum Roberts, Lewam Stefanos, Eric Fan-Lou, Bradley Pearce, Zachary T. Martin, Jose Medina Inojosa, Kristen Harris, Taha Ahmed, Alexander Razavi, Puja K. Mehta, J. Douglas Bremner, Paolo Raggi, Arshed Quyyumi

**Affiliations:** Department of Epidemiology, Rollins School of Public Health, Emory University, Atlanta, GA, United States; Department of Medicine, Division of Cardiology, Emory University School of Medicine, Atlanta, GA, United States; Veterans Administration Medical Center, Decatur, Georgia, United States; Department of Biostatistics and Bioinformatics, Rollins School of Public Health, Emory University, Atlanta, GA, United States; Department of Exercise Physiology, College of Health Sciences and Professions, Ohio University, Athens, OH, USA; Diabetes Institute, Ohio University, Athens, OH, USA; The Institute to Advance Population Health (ADVANCE), Ohio University, Athens, OH, USA; Department of Psychiatry and Behavioral Sciences, Emory University School of Medicine, Atlanta, GA, United States; Department of Radiology, Emory University School of Medicine, Atlanta, GA; Department of Medicine, Division of Cardiology, University of Alberta, Edmonton, Alberta, Canada

**Author notes:** **Address for Correspondence:** Viola Vaccarino, MD, PhD, Department of Epidemiology, Rollins School of Public Health, Emory University, 1518 Clifton Rd, Atlanta, GA 30329, USA.

## Abstract

**Background:** Women with premature myocardial infarction (MI) have worse outcomes than similarly aged men despite less severe disease, and stress-related factors have been implicated. We examined sex differences in the association of mental stress-induced myocardial ischemia (MSIMI) with cardiovascular outcomes after premature MI and explored vascular and inflammatory mechanisms.

**Methods:** We studied 602 individuals (mean age 51 years; 46% women) within 8 months of MI who underwent 99mTc-sestamibi myocardial perfusion imaging at rest and after mental stress. MSIMI was defined as a summed difference score ≥3. We measured reactive hyperemia index (RHI) as peripheral microvascular reactivity to stress and interleukin-6 (IL-6) as inflammatory response to stress. The primary endpoint was cardiovascular death or nonfatal MI; the secondary endpoint added heart failure hospitalization. Cox proportional hazards models adjusted for sociodemographic factors, clinical, and psychiatric history.

**Results:** MSIMI was more common in women than men (19.2% vs. 13.2%, p=0.04). Over a median follow-up of 34.2 months, 51 primary and 91 secondary endpoint events occurred. The hazard ratio (HR) of MSIMI for the primary endpoint was 3.2 (95% CI, 1.3, 7.6) in women and 1.9 (95% CI, 0.8, 4.5) in men (p for interaction=0.41), which remained similar after multivariable adjustment. The adjusted population attributable fraction was 27.6% (99% CI, 3.9%, 36.9%) in women and 11.1% (95% CI, −5.8%, 18.4%) in men. In exploratory analyses, all MSIMI-related primary endpoint events among women occurred in Black women (HR, 4.3; 95% CI, 1.7–11.1). Associations with the secondary endpoint were weaker and attenuated after adjustment. IL-6 and RHI responses did not attenuate associations.

**Conclusions:** Among women, MSIMI was associated with about threefold increased risk and over one quarter attributable fraction for cardiovascular death or nonfatal MI. Black women were at especially high risk. While larger studies are warranted to confirm sex differences and explore mechanisms, these data underscore the impact of emotional factors on morbidity and mortality in post-MI women.

## Introduction

In the U.S. and globally, coronary heart disease (CHD) is the number one killer of women,^1,2^ but despite scientific advances, it is unclear whether the pathophysiology of CHD differs between women and men.^3^ The study of CHD in women has historically been centered on older women. Yet women with premature myocardial infarction (MI), typically defined as an MI that occurs before age 65 in women, have emerged as a group in need of special study.^4–6^ Approximately 80,000 women are hospitalized for premature MI each year in the United States,^7^ and the incidence of hospitalization for premature MI, as well as the proportion of MI deaths occurring at younger ages, has increased among women in recent decades.^8,9^ We and others reported that this group has higher morbidity and mortality compared with men of similar age despite less severe disease.^10–13^ These disparities remain unexplained and suggest that there are sex differences in the pathophysiology, risk factors and prognostic factors of acute MI.

The psychosocial sphere is a largely neglected area for CHD prevention in women. We assembled a cohort of young and middle-aged individuals who survived an MI within the previous 8 months, with a large (>40%) representation of women—the Myocardial Infarction and Mental Stress study (MIMS). We found that social and emotional exposures mostly beginning early in life (such as depression, early life adversities, poverty) are more common in younger women with MI compared with men and community controls.^14^

In addition to being more prevalent, it is possible that emotional stress is a stronger risk factor in young women post-MI than other groups.^14–16^ A significant challenge, however, is to measure stress in a valid way. We used a standardized experimental approach to study mental stress-induced myocardial ischemia (MSIMI), a marker of stress vulnerability and significant prognostic factor in people with CHD.^5,17,18^ We found that younger women with MI had a higher occurrence of MSIMI than men of similar age.^14^ We also found that Black women are the group with the highest rate of MSIMI.^19^ Furthermore, our data have suggested that the mechanisms of MSIMI differ by sex. In men, but not in women, MSIMI is related to severity of angiographic stenoses^20^ and supply-demand mismatch from increased hemodynamic workload.^21^ In contrast, in women MSIMI is driven by microvascular vasoconstriction,^14,21^ a factor that is in itself related to adverse outcomes among women.^22^ Women with CHD, especially younger women, also have a more pronounced inflammatory response to stress which is also associated with adverse events,^23,24^ and therefore could contribute to adverse consequences in the setting of MSIMI.

Despite significant progress, many questions remain. First, the prognostic significance of MSIMI in women after MI is unknown, since former studies have mostly examined men, older people and cohorts with chronic CHD with insufficient inclusion of middle-aged women with a recent MI.^18,25^ Second, the pathways through which MSIMI might contribute to adverse events in post-MI patients are unclear. In women, in particular, processes involving vascular and immune function (especially stress-induced changes), could be especially important,^22,24^ but this has not been tested in survivors of a premature MI.

The current paper sought to examine the association of MSIMI with cardiovascular outcomes in young and middle-aged post-MI women in contrast to men of similar age and clinical condition, and to examine potential inflammatory and vascular mechanisms contributing to adverse outcomes related to MSIMI. We hypothesized that MSIMI is associated with adverse outcomes in women and that inflammation and vascular function are implicated.

## METHODS

Data for this analysis come from the second and third waves of the Myocardial Infarction and Mental Stress (MIMS) prospective cohort study.^14,19^ Wave 2 and Wave 3 were conducted using comparable methods following an earlier pilot phase (Wave 1).^26^ Wave 2 enrolled 313 participants between April 2012 and March 2017, while Wave 3 enrolled 306 participants between February 2020 and December 2023. Study protocols and laboratory procedures did not materially change between waves, including during the COVID-19 pandemic.

Participants were drawn from hospitals affiliated with Emory University across the Atlanta metropolitan area. To be eligible, individuals had to have a documented hospitalization for Type 1 MI within the preceding 8 months and to be 18-60 years old at the time of their MI hospitalization. Type 1 MI was verified through chart review according to standard diagnostic criteria, including elevated troponin together with ischemic symptoms, electrocardiographic changes, or other evidence of acute myocardial ischemia; obstructive coronary artery disease (CAD) was not a requirement for entry.^27^ Individuals were excluded if they had a serious co-existing medical or psychiatric illness likely to hinder follow-up or confound the results (e.g., cancer, renal failure, poorly controlled hypertension, ongoing alcohol or substance misuse, schizophrenia); were pregnant or nursing; or were taking immunosuppressants or psychotropic drugs other than antidepressants. Additional exclusions applied to those with unstable angina, acute MI, or decompensated heart failure in the week before enrollment; those weighing more than 450 pounds (a limitation of the nuclear imaging equipment); and those for whom study cardiologists judged it unsafe to hold anti-ischemic medications for the required 24-hour pre-testing period.

All participants completed a standardized mental stress protocol paired with myocardial perfusion imaging via single-photon emission computed tomography (SPECT), following established procedures.^28^ The Emory University Institutional Review Board approved all study procedures, and informed consent was obtained in writing from every participant.

### Mental Stress Testing

Following a 30-minute rest period, mental stress was elicited through a standardized public-speaking challenge.^14,28^ Participants were given a scenario involving emotionally charged content and asked to construct a believable narrative around it; they were told their speech would be judged on content, quality, and length. After two minutes of preparation, participants gave a three-minute talk before a video camera and three observers in white coats. Heart rate and blood pressure were captured every five minutes during the rest period and every minute during the speaking task. Perceived distress was measured with the Subjective Units of Distress Scale on a 0-100 linear rating.^29^

### Myocardial Perfusion Imaging

Each participant received two SPECT scans using Technetium-99m ([^99m^Tc]) sestamibi, one during rest and one during the stress task (with the tracer injected one minute into the speech). Weight-based dosing ranged from 10-14 mCi for the rest scan to 30-40 mCi for the stress scan. Both scans were performed the same day, with the stress scan scheduled at least 2 hours after the rest scan. Imaging occurred 45-60 minutes post-injection, consistent with published methods.^30^ ECG, heart rate, and blood pressure were monitored throughout, and anti-ischemic medications were held for the 24 hours preceding testing, per standard nuclear cardiology protocol. Scans were read visually by an experienced nuclear cardiologist (PR) blinded to participants’ demographic and clinical characteristics, using the standard 17-segment model of regional myocardial perfusion. Segments were rated on a 5-point scale (0 = normal, 1 = possibly normal, 2 = definitely abnormal, 3 = severely abnormal, 4 = no perfusion), from which summed stress, summed rest, and summed difference scores were derived.^30^ Consistent with prior work, mental stress-induced myocardial ischemia (MSIMI) was defined as a summed difference scores of 3 or greater.^31^

### Medical History and Other Clinical Information

Trained research personnel collected sociodemographic and medical history data and measured blood pressure as well as height and weight for BMI calculation. Race was self-reported. Data from medical records of the index MI were abstracted, including clinical details, procedures and angiographic findings on CAD severity assessed prior to any revascularization. Obstructive CAD was defined by ≥70% luminal stenosis, and disease severity was quantified using the Gensini score, which incorporates both the degree of stenosis and the prognostic importance of the affected coronary segment.^32^ Lifetime psychiatric diagnoses, including major depressive disorder and posttraumatic stress disorder (PTSD), were obtained via the Structured Clinical Interview for DSM-IV (SCID),^33^ administered by specially trained research personnel under the supervision of the study psychiatrist (JDB).

### Inflammation

Inflammatory markers were measured from venous blood samples using indwelling catheters collected at rest and 90-minutes post mental stress testing. Venous blood was collected in ice-cooled citrate tubes and immediately centrifuged at 4°C; plasma was frozen at -70°C until further processing. We employed the MesoScale multiplex assay system that uses electrochemiluminescence for high sensitivity and broad dynamic range. In this analysis, we focus on interleukin (IL)-6, a biomarker that acutely changes with mental stress,^34^ particularly among younger women,^23^ and is related to adverse outcomes in this group.^24^ The inter-assay coefficient of variations for midpoint standards for IL-6 was 5.8%, and the intra-assay coefficient of variation was 3.3%.

### Vascular Function

Peripheral microvascular function was assessed at rest and 30 minutes after the mental stress task. We measured the reactive hyperemia index (RHI) using the Endo-PAT2000 (Itamar Medical, Caesarea, Israel), which utilizes volume plethysmography to capture finger pulse volume amplitude (PVA), an indicator of peripheral blood volume change.^35,36^ PVA was recorded at rest and then during reactive hyperemia induced by inflating an upper-arm cuff to suprasystolic pressure for 5 minutes before release. A computer algorithm calculated RHI as the post-deflation-to-baseline PVA ratio in the cuffed-arm finger relative to the same ratio in the contralateral finger.^37^ Lower RHI values indicate greater peripheral microvascular dysfunction. The RHI was shown by our group and others to predict adverse cardiovascular outcomes in patients with CHD^38–40^ and among post-MI women.^22^

### Outcome Assessment

Participants were prospectively followed through annual contacts, with follow-up extending through May 31, 2026 (median follow-up 34.2 months). Study staff collected information on the incidence of cardiovascular events and hospitalizations through direct patient contact, medical record review, and by querying the vital records and the Social Security Death Index. If hospitalizations or procedures were reported, patients’ physicians were contacted, and hospital records were obtained. Events were adjudicated by study cardiologists unaware of other study data. The primary endpoint was a composite of cardiovascular death or nonfatal MI. We used the Type 1 MI definition of the Fourth Universal Definition of MI that included both obstructive and non-obstructive etiologies.^27,41^ A secondary endpoint added hospitalizations for heart failure.

### Statistical Analysis

We first described baseline demographic, behavioral and clinical characteristics in the overall sample and according to sex; no hypothesis testing was done for this descriptive comparison of baseline data.^42^ Next, we formally compared hemodynamic, vascular and inflammatory changes during mental stress between women and men, as well as occurrence of MSIMI and summed perfusion scores, using mixed-effects regression models with a random effect for study wave and adjusting for pretest values. In all models and for all statistical tests we used random effects to preserve the clustering within the two study waves. Crude incidence rates for the study endpoints were calculated as the number of events divided by total person-years of follow-up, in women and men and for MSIMI-positive and MSIMI-negative participants, and expressed per 100 person-years.

To assess the relationship between MSIMI and cardiovascular outcomes, we used Cox proportional hazards models with a frailty term following a gamma distribution to account for clustering by study. The proportional hazards assumption was assessed using Schoenfeld residual-based diagnostics and was satisfied. Sex-specific hazard ratios (HR) and 95% confidence intervals (CI) for MSIMI were derived by fitting an interaction term between MSIMI and sex. We performed multivariable analysis to adjust for sets of covariates deemed a priori potential confounding factors. Adjustment was conducted using sequential nested models to examine the effect of these adjustments on the model results. Model 1 was unadjusted; Model 2 was adjusted for sociodemographic and lifestyle characteristics (age, race, marital status, education >12 years, ever smoking, and BMI); Model 3 was further adjusted for clinical risk variables (history of hypertension, history of diabetes mellitus, left ventricular ejection fraction, and ST-segment elevation MI); Model 4 added lifetime history of major depression and PTSD and current use of antidepressant medications to determine whether results were driven by differences in psychiatric history. We derived sex-specific estimates, but since the interaction with sex was not significant in any of the models, we also reported the results in the overall sample including both women and men.

While the HR quantifies the relative strength of association between MSIMI and outcome, it does not capture the public health and clinical burden posed by MSIMI, which depends jointly on both the strength of association and the prevalence of MSIMI. To quantify this burden, we calculated the population attributable fraction (PAF), which is the proportion of outcome events attributable to MSIMI at the population level. Unadjusted and adjusted PAF were calculated using Miettinen’s formula: PAF = Pd(HR − 1) / HR,^43^ where Pd is the observed proportion of events occurring among MSIMI-positive participants and HR is the hazard ratio for MSIMI from Cox proportional hazards models. Given sparse events in some subgroups, 95% CIs for PAF were obtained by substituting the hazard ratio’s 95% CI bounds into the PAF formula, holding Pd fixed. PAF estimates with a lower confidence bound below zero were interpreted as not statistically significant.

Because of the high representation of Black women, we also examined results by sex and race. However, these analyses should be considered exploratory due to the small number of events in some subgroups, especially non-Black women.

In a separate set of models, we added inflammatory and microvascular responses to stress (change in IL-6 and RHI after mental stress) to examine whether adjustment for these stress responses attenuated the association of MSIMI with outcomes. No formal mediation analysis was conducted given that the initial results indicated no change in study estimates. IL-6 and RHI data were missing in approximately 20% of participants due to technical issues; missingness was not associated with sex, race, MSIMI status, or event rates. Therefore, this analysis was run separately in the subsamples with these data. For the main models, however, missing data were <5%; therefore, no imputation was conducted.

All analyses were conducted using SAS statistical software version 9.4 (SAS Inc., Cary, NC).

## RESULTS

### Study Sample and Baseline Characteristics

Complete imaging data were available for 602 of the 619 participants enrolled across the two MIMS waves (306 from Wave 2, 296 from Wave 3); the remaining 17 participants (7 from Wave 2, 10 from Wave 3) did not have imaging data because of technical issues with equipment or image acquisition unrelated to their health status. These 602 participants formed the analytic sample for this study (**Supplemental Figure 1**). The sample included 276 women (46%) and 326 men. Sample characteristics have already been described.^19^ Most participants identified as Black/African American (59%) or White (34%). Participants were, on average, middle-aged, with a mean age of 51 years (SD, 6.8 years, range, 25-61 years), with 93% falling between 40 and 60 years of age. All participants were ≤60 years old at the time of their MI, but 2 participants turned 61 years after their MI and before study enrollment. Time elapsed since the index MI averaged 4 months (with a median also of 4 months), a figure that held consistent in women and men.

Compared with men, women were more often Black, unmarried, and had lower income despite similar levels of education (**Table 1**). Cardiometabolic risk factors (BMI, history of diabetes and of hypertension) tended to be more elevated in women, and women also showed a higher prevalence of lifetime history of major depression and PTSD, but history of cardiovascular disease prior to the index MI was similar. Medication use was also overall similar, except that women were more likely to use antidepressants. At the index MI hospitalization, women had less ST-segment elevation MI compared with men, and their left ventricular ejection fraction was also higher, denoting a less clinically severe MI. Angiographic severity of CAD was also more favorable among women.

**Table 1.**
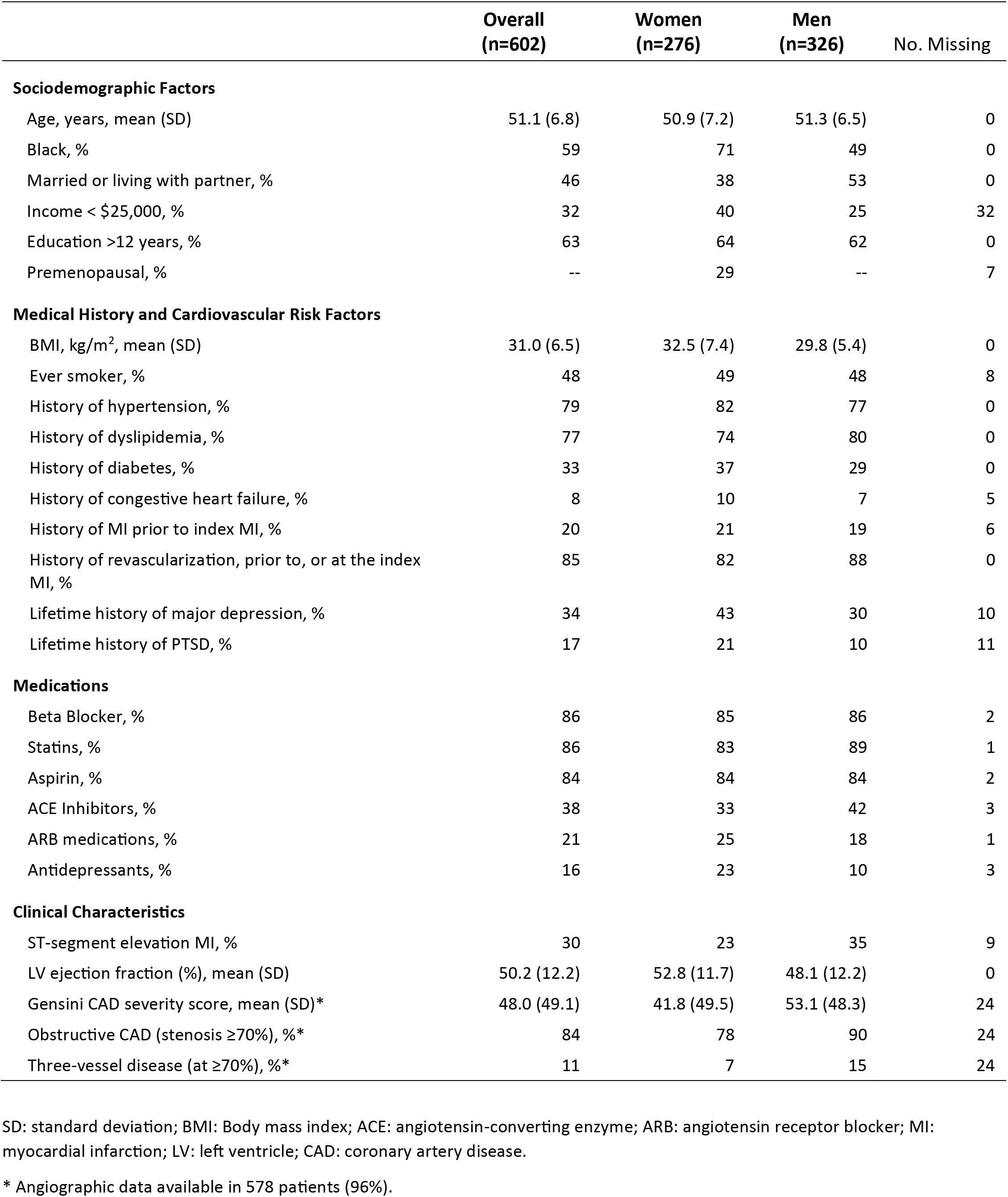
Characteristics of the study population by sex.

At rest, women had higher blood pressure and heart rate than men, but both groups showed a similar hemodynamic response to mental stress (**Table 2**). In contrast, women had both a higher resting IL-6 (denoting higher baseline inflammation) and a higher IL-6 increase with mental stress than men. Furthermore, women exhibited a lower resting RHI than men (denoting more microvascular endothelial dysfunction) which worsened with stress more in women than in men. Perceived distress was higher in women than men before and after mental stress.

**Table 2.**
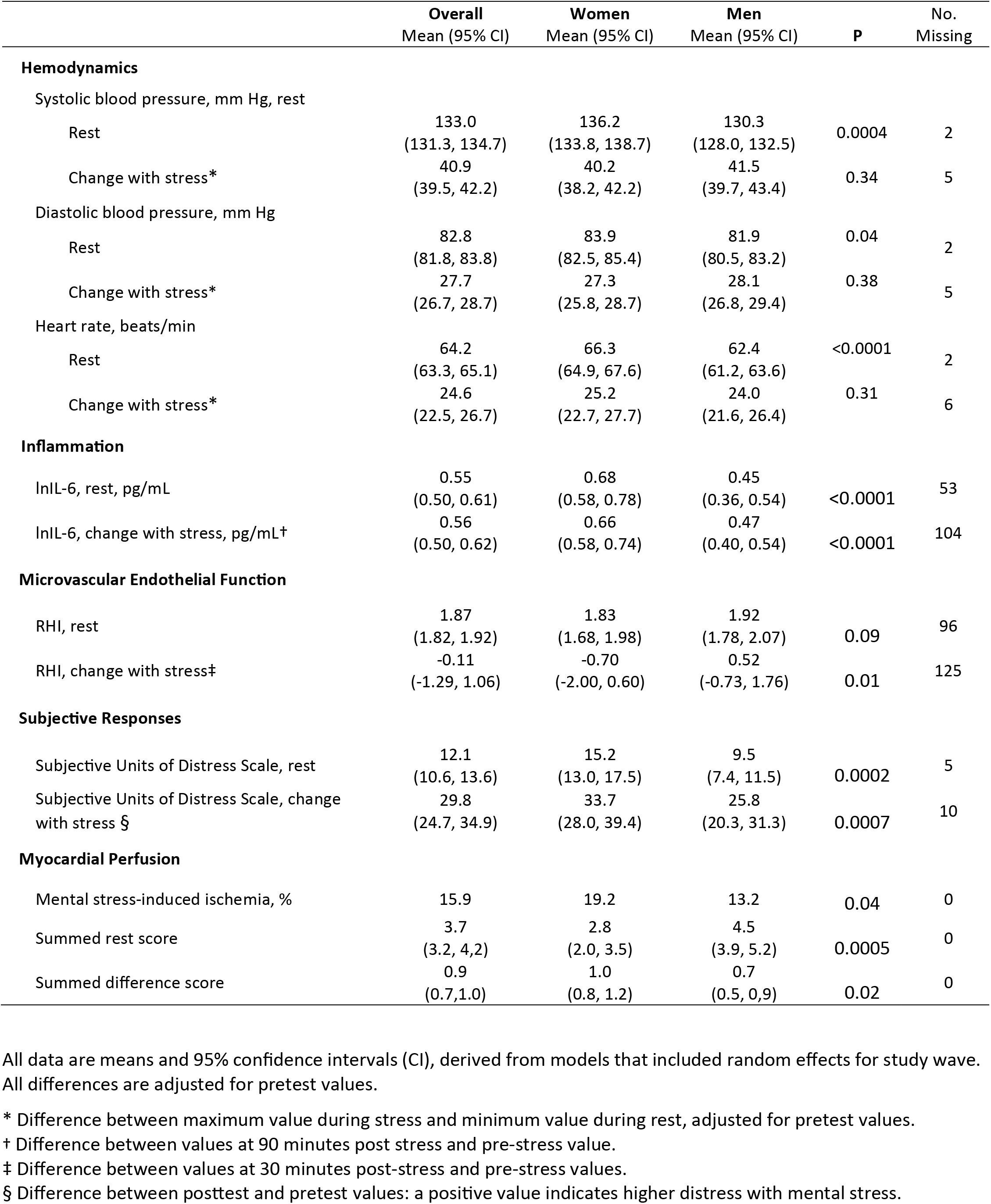
Differences in physiological and subjective responses to mental stress overall and by sex.

### Myocardial Perfusion

Overall, 96 (15.9%) of the participants developed MSIMI, with a similar rate in Wave 2 (16.3%) and Wave 3 (15.5%). Women were more likely to develop MSIMI than men: 19.2% vs. 13.2% (p=0.04) (**Table 2**). The summed rest score was significantly lower in women than in men (2.8 vs. 4.5, p<0.001), denoting fewer fixed (resting) defects, while the summed difference score was higher in women after adjusting for resting defects (1.0 vs. 0.7, p=0.02). Among women and men who developed MSIMI, however, both the summed rest score (4.8 in women and 4.4 in men, p=0.72) and the summed difference score (4.6 in women and 4.5 in men, p=0.70) were similar. These results suggest that, while women as a group showed more ischemic changes with mental stress than men (despite a lower prevalence of obstructive CAD), the severity of MSIMI was similar. As previously reported, MSIMI was most common among Black women, of whom 22% experienced MSIMI.^19^

### Association of MSIMI with Adverse Cardiovascular Events

The median follow-up time was 34.2 months (interquartile range [IQR], (24.9, 37.6 months), which was similar in women and men and in participants with MSIMI and without MSIMI (**Table 3**). Follow-up was completed in May 2026. Two participants could not be contacted at follow-up and therefore their follow-up time was censored shortly after the baseline visit.

**Table 3.**
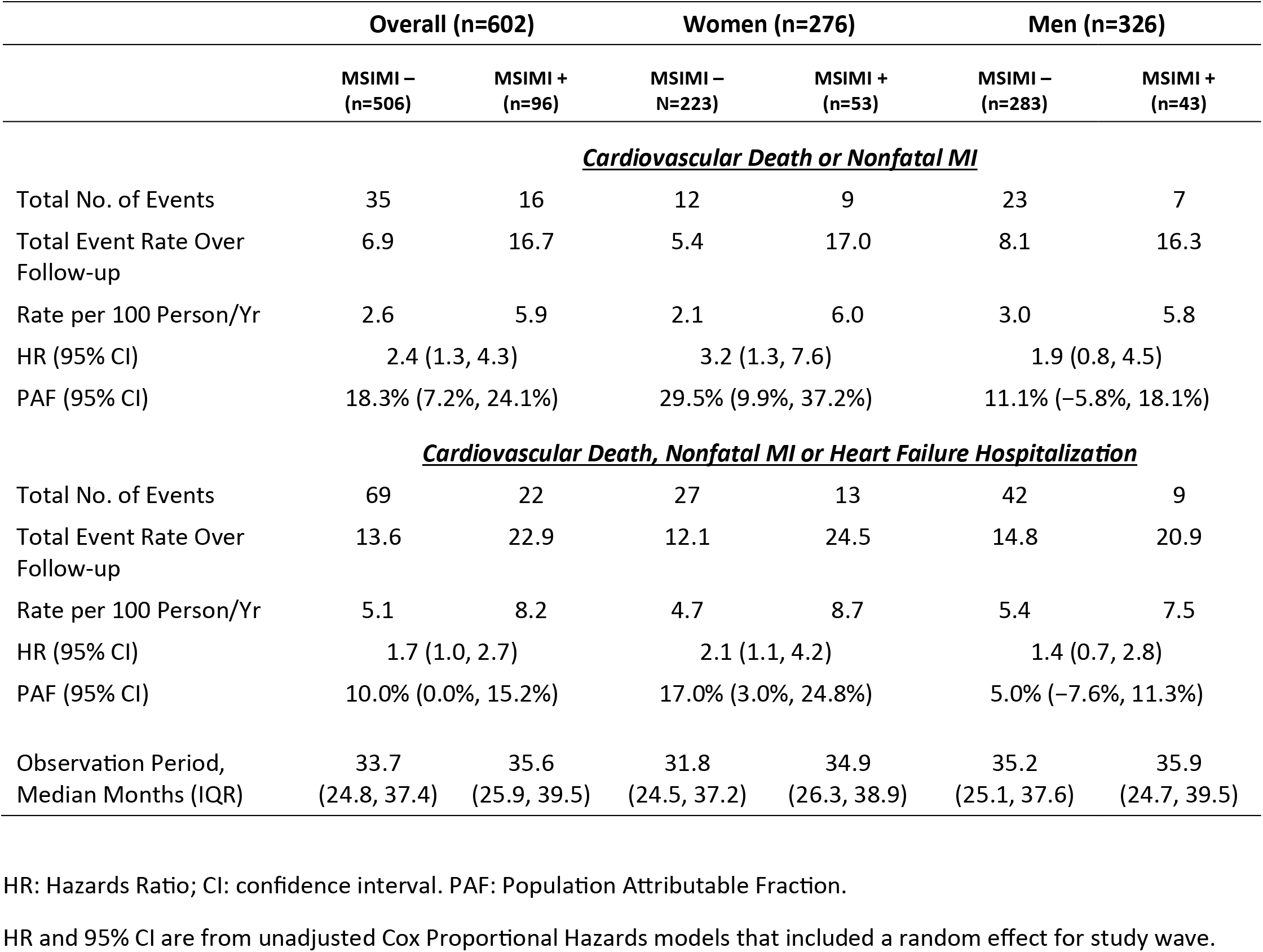
Incidence of Cardiovascular Outcomes by Mental Stress–Induced Myocardial Ischemia.

Over the follow up, there were a total of 51 events for the primary endpoint of cardiovascular death or nonfatal MI and 91 events for the secondary endpoint of cardiovascular death, nonfatal MI or hospitalizations for heart failure (**Table 3**). Both the primary and the secondary endpoint occurred more often in participants with MSIMI than in those without MSIMI. The event rate of the primary endpoint was 2.6 per 100 person-years for those without MSIMI and 5.9 per 100 person-years for those with MSIMI; the unadjusted HR comparing patients with MSIMI to those without MSIMI was 2.4 (95% CI, 1.3, 4.3). While the rate of events per 100 person-years was lower in women than in men without MSIMI, it surpassed the rate of men among those with MSIMI. In women, the HR for the primary endpoint was 3.2 (95% CI, 1.3, 7.6), and in men, the HR was 1.9 (95% CI, 0.8, 4.5). The unadjusted PAF in women was 29.5% (95% CI, 9.9%, 37.2%), while in men it was 11.1% (95% CI, −5.8, 18.1%). These estimates indicate that close to 30% of events in women was attributable to MSIMI, vs. 1 in 9 events in men (the latter not statistically significant).

The association with MSIMI was weaker for the secondary endpoint than for the primary endpoint (**Table 3**). Because the secondary endpoint includes the primary endpoint events, this attenuation implies little or no association of MSIMI with heart failure hospitalizations. For the secondary endpoint, the event rate was 5.1 per 100 person-years for patients without MSIMI, and 8.2 per 100 person-years for those with MSIMI, with an HR of 1.7 (95% CI, 1.0, 2.7). In women, the HR for the secondary endpoint was 2.1 (95% CI, 1.1, 4.2), and in men, the HR was 1.4 (95% CI, 0.7, 2.8). The PAF in women was 17.0% (95% CI, 3.0%, 24.8%) and in men it was 5.0% (95% CI, -7.6%, 11.3%).

In exploratory analyses by sex and race (**Supplemental Table 1**), among women all 9 MSIMI-related events for the primary endpoint occurred among Black women, with an HR for MSIMI in Black women of 4.3 (95% CI, 1.7, 11.1). Black men also had an elevated risk for the primary endpoint (HR, 3.0, 95% CI, 1.1, 8.6), while there was no association among non-Black men (HR, 1.0, 95% CI, 0.2, 4.6), p=0.06 for the interaction of MSIMI with race. For the secondary endpoint, all but one of the MSIMI-related events in women occurred among Black women, with an HR in this group of 2.1 (95% CI, 1.0, 4.1). Because of the small number of events, these subgroup estimates should be interpreted with caution.

Adjustment for demographic, clinical, and even psychiatric history did not materially change the association of MSIMI with cardiovascular outcomes (**Table 4**). In all models for the primary endpoint, the association was larger in women than in men and statistically significant in women only, with almost threefold increased risk among women, vs. about twofold increased risk among men, although the interaction of sex and MSIMI was not statistically significant (**Table 4**, **Figure 1**). The fully adjusted PAF (Model 4) held similar to the unadjusted estimate: overall, 17.7% (95% CI, 5.2%, 24.1%); in women, 27.6% (95% CI, 3.9%, 36.9%); and in men, 11.1% (95% CI, −5.8%, 18.4%). For the secondary endpoint, associations were attenuated after adjusting for clinical risk factors, and the fully adjusted estimate for MSIMI was no longer statistically significant in either women or men (**Table 4**).

**Figure 1.**
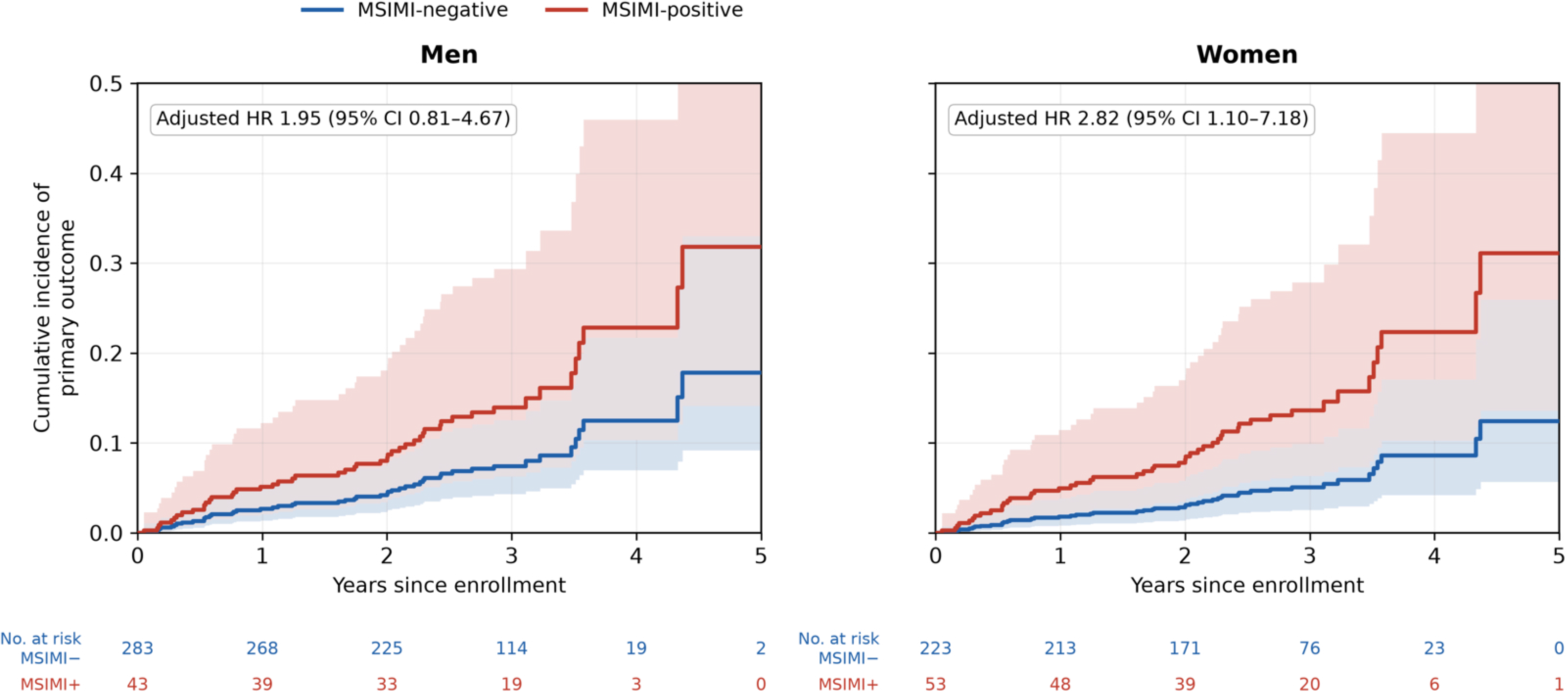
Adjusted Cumulative Incidence of the Primary Outcome (Cardiovascular Death or Nonfatal MI), by MSIMI Status and Sex. Graphs are derived from a pooled Cox model that was fully adjusted for Model 4 variables (age, race, marital status, education, ever smoking, BMI, hypertension, diabetes mellitus, left ventricular ejection fraction, ST-segment elevation MI, and DSM-IV diagnosis of depression and of PTSD).

**Table 4.** Association between mental stress induced myocardial ischemia and the study endpoints.

|  | Overall | Women | Men | P for<br>interaction<br>of sex and<br>MSIMI | No. in<br>Model |
| --- | --- | --- | --- | --- | --- |
|  | HR (95% CI) |  |  |  |  |
| ***Cardiovascular Death or Nonfatal MI*** |  |  |  |  |  |
| **Model 1:** Unadjusted | 2.4 (1.3, 4.3) | 3.2 (1.3, 7.6) | 1.9 (0.8, 4.5) | 0.41 | 602 |
| **Model 2:** Adjusted for sociodemographic and lifestyle factors* | 2.5 (1.3, 4.5) | 3.0 (1.3, 7.3) | 2.0 (0.8, 4.8) | 0.51 | 594 |
| **Model 3:** Adjusted for the above plus CAD risk factors and CAD severity indicators† | 2.3 (1.2, 4.2) | 2.7 (1.1, 6.9) | 1.9 (0.8, 4.6) | 0.60 | 585 |
| **Model 4:** Adjusted for the above plus lifetime history of depression and PTSD and current antidepressant use | 2.3 (1.2, 4.3) | 2.8 (1.1, 7.2) | 1.9 (0.8, 4.7) | 0.58 | 574 |
| ***Cardiovascular Death, Nonfatal MI or Heart Failure Hospitalization*** |  |  |  |  |  |
| **Model 1:** Unadjusted | 1.7 (1.0, 2.7) | 2.1 (1.1, 4.2) | 1.4 (0.7, 2.8) | 0.36 | 602 |
| **Model 2:** Adjusted for sociodemographic and lifestyle factors* | 1.9 (1.2, 3.1) | 2.1 (1.1, 4.3) | 1.6 (0.8, 3.4) | 0.55 | 594 |
| **Model 3:** Adjusted for the above plus CAD risk factors and CAD severity indicators† | 1.5 (0.9, 2.5) | 1.5 (0.7, 3.0) | 1.5 (0.7, 3.1) | 0.97 | 585 |
| **Model 4:** Adjusted for the above plus lifetime history of depression and PTSD and current antidepressant use | 1.6 (0.97, 2.7) | 1.7 (0.8, 3.6) | 1.5 (0.7, 3.2) | 0.78 | 574 |

HR: Hazards Ratio; CI: confidence interval; CAD: coronary artery disease; PTSD: posttraumatic stress disorder;. HR and 95% CI are from Cox proportional hazards models that included a random effect for study wave.
\*Age, race (black versus non-black), marital status (married/living with partner vs not), education >12 years, ever smoking, and BMI.
† Hypertension, diabetes mellitus, left ventricular ejection fraction, and ST-segment elevation MI.

As shown in **Table 5**, data on IL-6 before and after mental stress were available in 498 participants (82.7%), and data on RHI were available in 477 (79.2%). Unadjusted results were similar in these subsamples compared with the main sample, even though, as expected given the reduced sample size, confidence intervals were in general wider. After adjusting for changes in IL-6 and RHI with mental stress, respectively, results remained unchanged for both endpoints, suggesting that inflammatory and peripheral microvascular responses to stress did not contribute to the association, either in the overall sample or when women and men were examined separately.

**Table 5.**
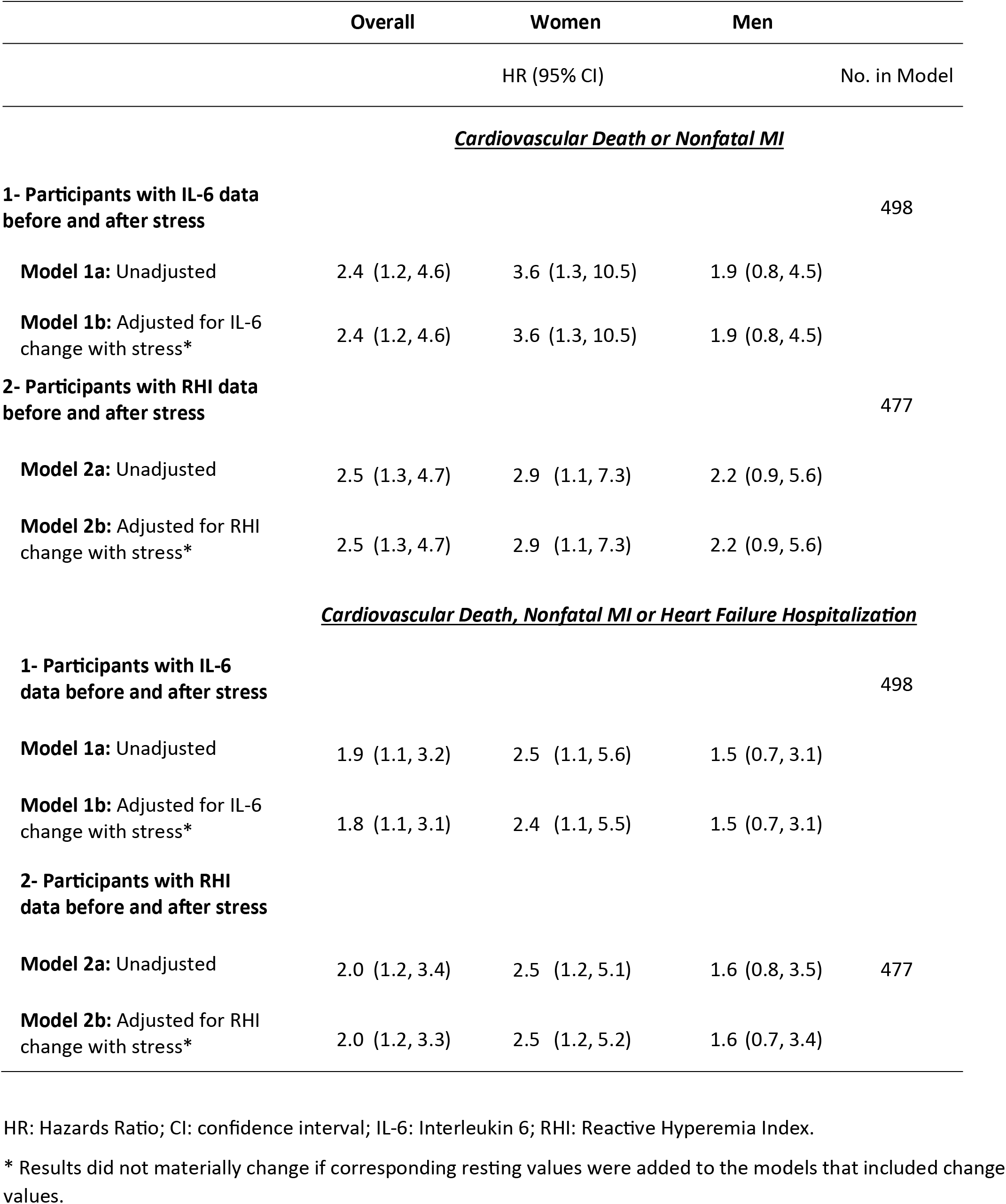
Association between mental stress induced myocardial ischemia and the study endpoints after adding inflammation and vascular function variables.

## DISCUSSION

Among young and middle-aged survivors of an MI, women who developed MSIMI assessed with myocardial perfusion imaging had approximately threefold higher risk of cardiovascular death or nonfatal MI compared with women who did not develop MSIMI after a median follow-up of 34.2 months. Over one quarter (27.6%) of primary outcome events, including cardiovascular death or nonfatal MI, could be attributable to MSIMI among women, which was 2.5 times the attributable fraction for men (11.1%). The risk associated with MSIMI was highest among Black women, but also significantly elevated among Black men, although race-stratified analyses were exploratory. In the overall sample and among women, the association was stronger for the primary endpoint of cardiovascular death or nonfatal MI, while it was attenuated when hospitalizations for heart failure were included in the composite outcome. Inflammatory and vascular responses to stress were more pronounced among women, but contrary to our expectations, they did not help explain the association between MSIMI and outcomes, either among women or in the overall sample.

The interaction between MSIMI and sex was not statistically significant, so the apparent difference by sex for the primary endpoint should be interpreted with caution and further evaluated in larger studies. Nonetheless, the magnitude of the MSIMI-associated risk in women is consistent with a growing body of evidence that women with CHD, especially younger women, are more vulnerable to the cardiovascular effects of psychological stress, including a higher propensity to develop MSIMI, more pronounced microvascular and inflammatory responses to stress, and stronger associations of these responses with adverse outcomes.^16–21,26–29^

Women had higher rates of MSIMI than men, a finding that has already been reported.^14,16,26,44^ The prognostic impact of MSIMI in patients with CHD is also well established.^18,25^ However, previous studies have included a limited number of women, especially women with a premature MI, a growing group with higher morbidity and mortality compared with men of similar age despite that the women have less severe disease.^10–13^ Our own prior work demonstrated that MSIMI predicts adverse cardiovascular events in predominantly older patients with chronic CHD,^25^ but that study was not designed to examine outcomes specifically in women, nor in a population enriched for a recent, premature MI. To our knowledge, the present study is the first to examine the prospective association between MSIMI and hard cardiovascular outcomes in a cohort of young and middle-aged survivors of a premature MI with a large representation of women, and to suggest that this risk may be concentrated among Black women. Because of the high rate of MSIMI in younger post-MI women, and its strong relationship with adverse events, the clinical impact of MSIMI in this group may be large: the estimated PAF of 27.6% suggests that, assuming the association is causal and unconfounded, nearly 30% of adverse events in this group could be attributable to MSIMI.

Despite comprehensive measurements spanning behavioral and biological variables, we were unable to elucidate the mechanisms underlying the excess risk associated with MSIMI in post-MI women. Hemodynamic responses to mental stress are unlikely to explain the sex differences in risk associated with MSIMI. There were no differences in blood pressure and heart rate changes with mental stress between women and men; furthermore, such responses are not important correlates of MSIMI among women.^21^ Severity of ischemia also cannot explain the results, since there were no differences in the severity of perfusion abnormalities between women and men with MSIMI. Socioeconomic position is an established driver of cardiovascular health, primarily through lifestyle and worsening of cardiovascular risk factors,^45,46^ and socioeconomic disadvantage affects mortality after MI in younger patient populations.^47^ However, in our analysis adjustment for socioeconomic and cardiovascular risk factors did not explain the excess risk associated with MSIMI in women. Based on previous work,^22,24^ we had hypothesized that inflammatory and vascular responses to stress, which were more unfavorable among women, would be implicated. However, these responses did not explain the risk associated with MSIMI among women. Taken together, the persistence of the association between MSIMI and outcome despite adjustment for these established pathways suggests that other unmeasured biological or psychosocial mechanisms may be responsible, and warrant investigation in future studies.

Virtually all adverse events in women with MSIMI occurred among Black women, a group that in the U.S. faces significant health burdens and higher case fatality after a premature MI.^8,48^ Possible explanations for our findings include the higher prevalence of cardiometabolic comorbidities in this subgroup which may not have been fully accounted for in our analysis, socioeconomic disadvantage, and differential access to post-MI care, but also the small number of events among non-Black women, which limits the precision of these comparisons. Furthermore, MSIMI was associated with elevated risk also among Black men, suggesting that some of the relevant mechanisms may be shared among Black individuals rather than being specific to women. More work is needed in larger studies with high representation of Black post-MI individuals.

The risk associated with MSIMI was mostly related to subsequent ischemic events (recurrent MI or cardiovascular death) rather than heart failure. This is not surprising given that MSIMI operates through acute, transient physiologic perturbations driven by sympathetic surge and vagal withdrawal, including catecholamine release, coronary and peripheral microvascular constriction and platelet activation.^17,49,50^ These same pathways are implicated in plaque rupture/erosion, coronary vasospasm, and microvascular thrombosis which may trigger recurrent MI and ischemic/arrhythmic cardiovascular death. Heart failure progression, by contrast, is driven largely by structural and hemodynamic processes such as infarct size, left ventricular remodeling and chronic neurohormonal activation, pathways less likely to be altered by episodic ischemic triggers. Furthermore, there is evidence that mental stress may facilitate lethal ventricular arrhythmias, a pathway to cardiovascular death that bypasses heart failure progression entirely.^51^

Our findings have substantial implications. The incidence of hospitalization for premature MI has increased among women,^9^ and premature MI accounts for a growing proportion of MI deaths among women and especially Black women.^8^ Despite considerable progress in improving the awareness of cardiovascular disease as a major cause of morbidity and mortality in women, such awareness is still low, particularly among younger women and minority women.^52,53^ The high risk associated with MSIMI in post-MI midlife women necessitates a renewed focus on stress-related risk pathways for this vulnerable population. Most Class 1 secondary prevention recommendations for patients with chronic CHD emphasize traditional risk factors and antiplatelet therapy, with little guidance for psychosocial factors; screening and management for mental health currently carries a Class 2a recommendation.^54^ Our findings highlight the need for targeted risk assessment protocols and prevention strategies specifically designed to address the unique risks of women post-MI. For example, risk stratification algorithms could be devised that include psychological assessments. Wearable devices or smartphone apps could be employed to monitor real-time physiological and perceived responses to stress and heart rate variability patterns,^55^ especially given that these measures, taken during everyday life in post-MI patients, are more unfavorable among women.^56^ The effectiveness of these approaches, however, requires future evaluation.

Our study is strengthened by a large representation of women, the assessment of MSIMI using myocardial perfusion imaging, a well-validated and standardized modality for ischemia detection,^30^ and rigorous methodology in the assessment of events at follow-up, which were independently adjudicated. To our knowledge, this is the largest follow-up study to date of young and middle-aged survivors of an MI who underwent assessment of MSIMI. Even so, the number of events was small in some subgroups, which limited the ability to test interaction effects. The women in our study were primarily Black or African American. There were only 81 women who were not Black, and these women only had a total of 4 and 5 events for the primary and secondary endpoint, respectively. Therefore, our study cannot provide insights for women with race/ethnicity other than Black. Our study used a single mental stressor, as opposed to multiple triggers. Even though some consider this a limitation, others have argued that using multiple stressors in sequence may cause undesirable cumulative effects.^57^ A single stressor is necessary when using myocardial perfusion imaging with ^99m^Tc, which provides a “snapshot” of perfusion during the stressful task, with the advantage that the patient does not need to lie in a scanner during the test. We cannot exclude selection bias due to potential exclusion of MI survivors who were too sick or otherwise unable to participate. Finally, our study population was drawn from hospitals in a single city and conducted at a single institution. The generalizability of our findings to other populations and settings requires further exploration.

In conclusion, MSIMI identifies a subgroup of midlife post-MI women, particularly Black women, at markedly elevated risk of recurrent MI and cardiovascular death through pathways that remain to be discovered. These findings argue for greater clinical attention to psychological stress reactivity as a potentially modifiable, or at least identifiable, risk marker in the care of women after MI.

## Data Availability

Data will be available from the corresponding author upon reasonable request.

## Supplemental Material

Supplemental Table 1, Supplemental Figure 1.

